# The Ecological Structure of Mosquito Population Seasonal Dynamics

**DOI:** 10.1101/2021.01.09.21249456

**Authors:** Charles Whittaker, Peter Winskill, Marianne Sinka, Samuel Pironon, Claire Massey, Daniel J Weiss, Michele Nguyen, Peter W Gething, Ashwani Kumar, Azra Ghani, Samir Bhatt

## Abstract

Understanding the temporal dynamics of mosquito populations underlying malaria transmission is key to optimising control strategies. We collate mosquito time-series catch data spanning 40 years and 117 locations across India to understand the factors driving these dynamics. Our analyses reveal pronounced variation in dynamics across locations and between species. Many mosquito populations lacked the often-assumed positive relationship with rainfall, instead displaying patterns of abundance that were only weakly or even negatively correlated with precipitation and highlighting the role of temperature, proximity to perennial bodies of water and patterns of land use in shaping the dynamics and seasonality of mosquito populations. We show that these diverse dynamics can be clustered into “dynamical archetypes”, each characterised by distinct temporal properties and driven by a largely unique set of environmental factors. These results highlight that a complex interplay of factors, rather than rainfall alone, shape the timing and extent of mosquito population seasonality.

## Background

With an estimated 229 million cases and over 400,000 deaths across 87 endemic countries in 2019^1^, malaria represents one of the most serious infectious diseases globally^2^. Nineteen countries in sub-Saharan Africa along with India account for almost 85% of the global burden^3^, with *Plasmodium falciparum* most prevalent in African settings, and India alone accounting for almost 50% of the global *Plasmodium vivax* burden^4^. Transmission occurs via mosquito vectors belonging to the *Anopheles* genus – these vectors are heterogeneously distributed across the globe^5,6^, a feature that results in marked differences in the transmission dynamics of malaria across different ecological contexts.

Much work has focussed on characterising the global spatial distribution (presence/absence) of these malaria vectors^7,8^. This work represents a vital input to surveillance and control programmes aimed at mitigating the impacts of vector borne diseases worldwide. By contrast, less attention has been paid to understanding the temporal patterns of vector abundance, and how these dynamics are shaped by the local environment. Mosquito populations are highly temporally dynamic, exhibiting substantial annual fluctuations in size^9,10^ that drive the temporal profile of malaria risk. Understanding the determinants of these dynamics is important given that the efficacy of many malaria control interventions (such as seasonal malaria chemoprevention^11,12^ and indoor-residual spraying^13,14^) depends on the timing of their delivery in relation to seasonal peaks in malaria risk. Effective utilisation of these interventions will be vital for achieving the goals of the World Health Organisation*’*s “High Burden, High Impact” strategy, which aims to substantially reduce/eliminate malaria in India and the ten African nations with the highest global burden^15^.

Despite their importance, many questions remain surrounding the drivers of mosquito population dynamics. Rainfall is frequently considered a key determinant of mosquito temporal dynamics due to the requirement of an aquatic habitat for the early life cycle stages, with many species displaying a preference for transient, rain-fed pools of water in which to breed^16^. However, whilst a close relationship has been observed between rainfall occurrence, peaks in mosquito populations and malaria cases^17^ (e.g. *Anopheles gambiae s.l.*^18–20^ for African settings and *Anopheles dirus s.l.* across India and south-East Asia^21^), *Anopheles funestus s.l.* and *Anopheles annularis s.l.* populations frequently lack marked seasonal fluctuations in population abundance^22,23 10,24,25^. This brings into question how generalisable relationships between rainfall and mosquito population dynamics are. The influence of other factors such as temperature (which has a marked influence on many mosquito traits including larval development^26^, biting rates and mortality rates^27^) remains similarly unclear. Recent field-based work has suggested that considerations of both rainfall and temperature are necessary to understand seasonal patterns of malaria incidence^28^. However, these analyses have been restricted to a small number of settings across sub-Saharan Africa; leaving the influence of temperature regimen on mosquito population dynamics largely unexplored in other ecological settings.

Altogether, these results highlight outstanding questions surrounding the drivers of mosquito population dynamics. Using India as a case study, we collate a dataset of temporally disaggregated mosquito catch data from across the country to better understand variation in mosquito population dynamics and the factors underlying this variation. We use these data to characterise the temporal patterns displayed by different mosquito species complexes and identify pronounced heterogeneity in the extent and nature of seasonal dynamics, both between species complexes and across different locations. Exploring the drivers of these dynamics highlights the critical importance of both abiotic and species-specific factors in shaping temporal patterns of mosquito abundance. It also underscores the importance of considering both species composition and ecological structure when implementing malaria control interventions.

## Results

### Substantial Diversity in Mosquito Population Dynamics Within and Between Species

A total of 272 time-series from 117 locations across India were identified through the systematic review, spanning seven species complexes that together represent the dominant malaria vectors in the country **(Fig.1A)**. These time-series were then smoothed using a Negative Binomial Gaussian Process based framework **(Fig.1B)**. Substantial variation in temporal dynamics was observed between different species complexes with many of the collated time-series lacking the close, positive correlation with rainfall typically assumed for mosquito populations. Whilst *Anopheles dirus s.l.* populations tended to peak during the monsoon period (typically June to September), many *Anopheles fluviatilis s.l.* populations by contrast peaked between November and February (the dry season across most of India), reaching their lowest density during the monsoon. Despite highly seasonal patterns of rainfall, a number of time-series belonging to *Anopheles annularis s.l.* demonstrated perennial patterns of abundance. In addition to this variation between species complexes, we also observed extensive variation in temporal dynamics within a species complex. Across the 85 time-series collated for *Anopheles culicifacies s.l.*, populations varied substantially in both the extent and timing of their seasonal peaks; this ranged from sharp peaks in the monsoon season to perennial characteristics more similar to those observed for *Anopheles annularis s.l.*. A range of dynamics were also observed for time-series belonging to *Anopheles stephensi s.l.*, from peaks coincident with the monsoon season to bimodal dynamics displaying peaks both during and outside the rainy season.

**Figure 1:**
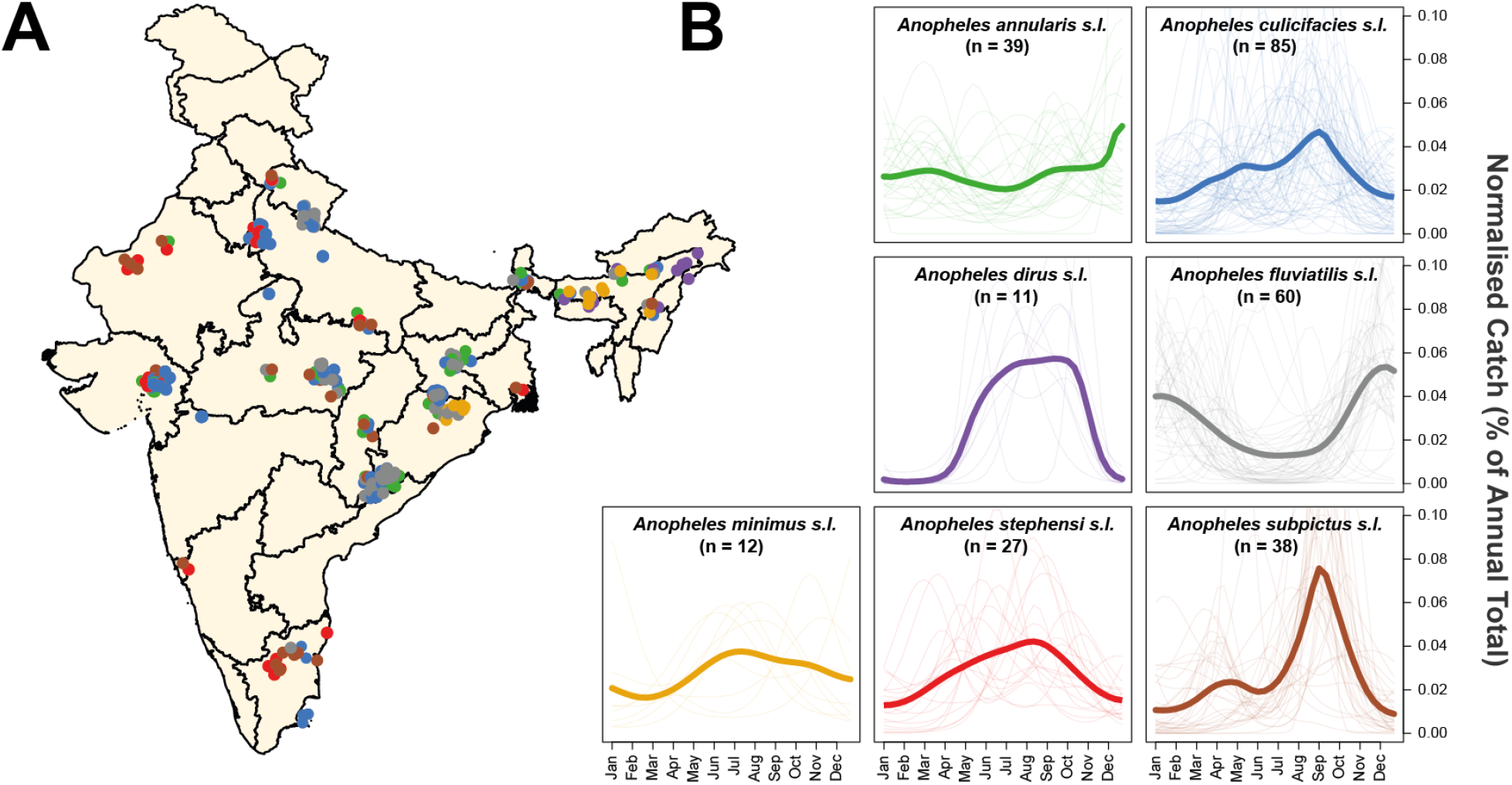
Exploring Species Complex-Specific Patterns of Mosquito Population Dynamics. Negative Binomial Gaussian Processes incorporating a periodic kernel were fitted to each of the 272 time-series collected from 118 locations across India collated as part of the systematic review. These fitted time-series (representing monthly catches over the course of a year) were then normalised and the results plotted here, disaggregated by species complex. **(A)** Map of India showing the different locations for which time-series data was available. Points represent a single collected time-series, coloured according to the species complex. **(B)** Normalised, Gaussian Process fitted time-series disaggregated by species complex. In all instances, pale lines represent a single time-series for that particular species complex, and the brighter line is the mean of all of the time-series belonging to that species complex, evaluated at that particular timepoint.

### Statistical Characterisation of Mosquito Catch Time-Series Properties Reveals Distinct Temporal Patterns

An array of summary statistics were calculated for each time series in order to characterise their temporal properties (see **Supplementary Methods** and **Supp Fig.2** for more information). This was followed by k-means clustering of the results, to assess whether the observed variation could be delineated into discrete groups, each characterised by distinct temporal patterns. We identified 4 groups (**Fig.2A)** – these included time-series peaking during the monsoon season (Cluster 1), displaying bimodal characteristics (Cluster 2), peaking in the dry season (Cluster 3) or displaying perennial patterns of abundance (Cluster 4) **(Fig.2B)**. Cluster assignment was robust to the choice of prior used in the time-series fitting and smoothing (**Supp Fig.3**). The distinct patterns displayed by each group were not due to differences in the timing and extent of rainfall across India – we observed a high positive cross-correlation product between rainfall and mosquito density for Cluster 1 (r=0.52), but a negative correlation for Cluster 3 (r=-0.41) and low correlation for Clusters 2 and 4 (r=-0.08 and 0.03 respectively, **Supp Fig.4**). This suggests that the observed patterns represent genuine differences between species and across locations in how mosquito populations respond to rainfall.

**Figure 2:**
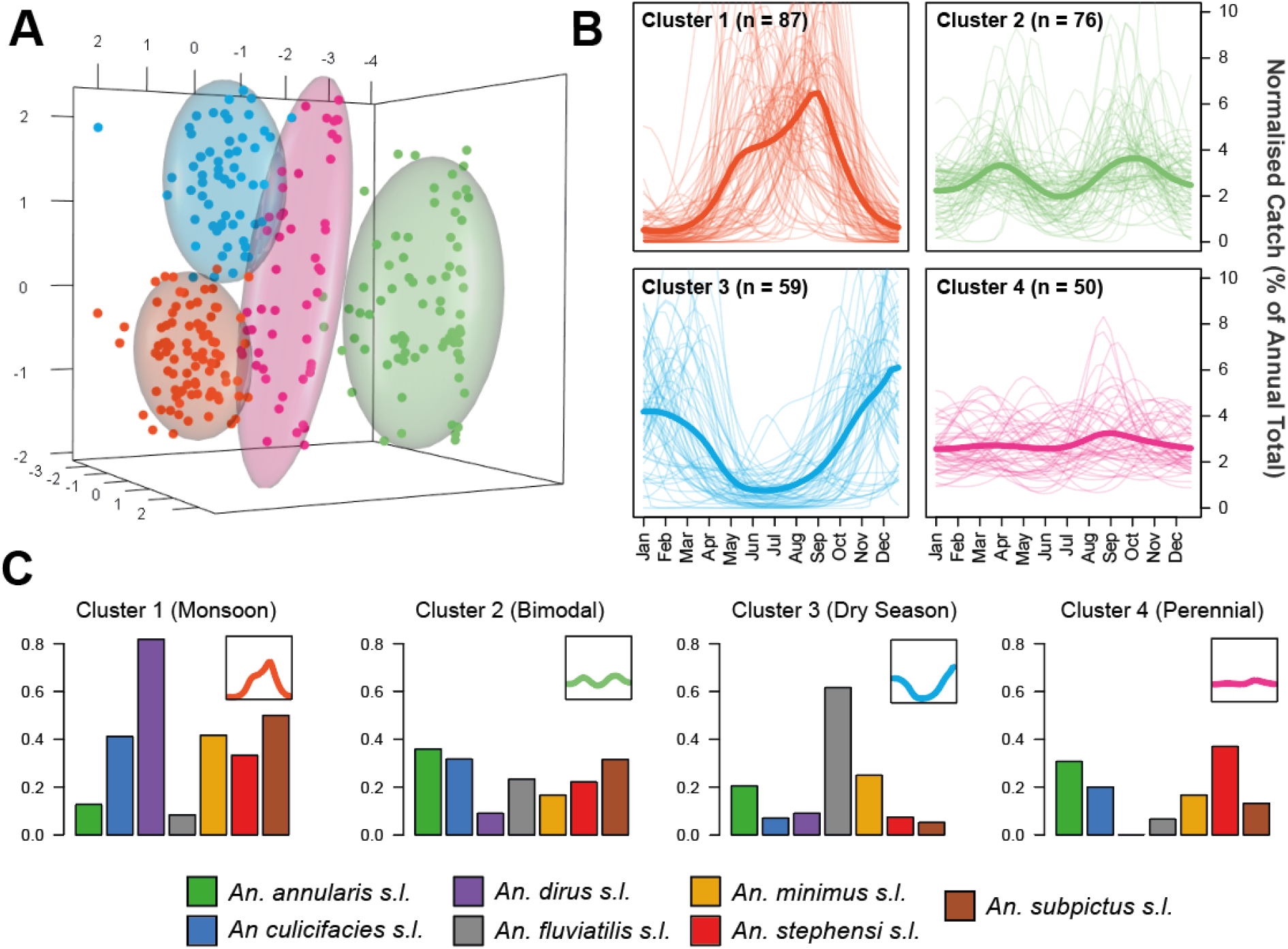
Characterisation and Clustering of Time-Series with Similar Temporal Properties. Statistical characterisation of the properties of each time series was followed by Principal Components Analysis and the results clustered using the k-means algorithm. **(A)** Results of the k-means clustering algorithm for 4 clusters, with a Principal Components Analysis applied for visualisation purposes. Colour of the points refers to cluster membership, coloured ellipsoids demarcate the 75^th^ quantile of the density associated with each cluster. First 3 principal components are plotted, explaining 53%, 15% and 14% of the overall variation, respectively. **(B)** The time-series belonging to each cluster. Pale lines represent individual time-series, brighter line represents the mean of all the time-series belonging to that cluster, evaluated at each timepoint. Characterisation and clustering in this way revealed distinct groups of time-series that share similar temporal properties. **(C) T**he proportion of time-series for each species complex belonging to each cluster - different coloured bars indicate different species complexes (see legend) and y axis corresponds to the proportion of time-series (for a given species complex) belonging to that cluster.

For some species complexes, the majority of their time-series belonged to a single cluster (**Fig.2C**) – *Anopheles dirus s.l* time-series were restricted primarily to Cluster 1 (monsoon season peaking) whilst *Anopheles fluviatilis s.l.* time-series were almost exclusively found in Cluster 3 (dry season peaking). By contrast, time series belonging to *Anopheles culicifacies s.l.* appeared across all four clusters – indicating that different sibling species within the complex display distinct temporal dynamics or that mosquito populations belonging to the species complex are able to adopt a diverse array of temporal dynamics depending on the particular ecological setting.

### Mosquito Population Dynamics are Determined by a Complex Interplay of Abiotic and Biotic Factors

Using binary indicators for species complex (seven total, indicating which species complex a particular time-series belongs to) and a suite of ecological variables (25 total) as predictors, we fitted a multinomial logistic regression model to the cluster labels (i.e. which cluster each time-series had been assigned to) to explore potential factors underlying the observed variation in temporal dynamics. This framework produces one coefficient estimate for each cluster and predictor (a total of 4 coefficients per cluster and predictor), with that coefficient defining the strength of the association between a predictor and a particular cluster.

Across the species complex regression coefficients, *Anopheles culicifacies sl.* and *Anopheles subpictus s.l.* demonstrated large and positive associations with Cluster 1 (monsoon peaking dynamics), whereas for *Anopheles fluviatilis s.l.*, this relationship was strongly negative (the species-complex associated with Cluster 3 instead). To explore this variation more systematically, we employed a hierarchical clustering approach to examine the coefficient values across all species-complexes simultaneously and identified significant structuring **(Fig.3A).** In contrast to *Anopheles culicifacies s.l.* and *Anopheles subpictus s.l.* (which clustered together and showed a strong positive association with Cluster 1 and a strong negative association with Cluster 3), the binary indicator variable for *Anopheles dirus s.l.* displayed weak associations with all clusters, including for Cluster 1 which the majority of the collated time series for this species-complex had been assigned to. This suggests that the ecological features of the locations *Anopheles dirus s.l.* had been sampled in (as measured by the environmental covariates), rather than features intrinsic to species-complex itself (as captured by the species binary indicator), were the primary driver of the observed dynamics.

**Figure 3:**
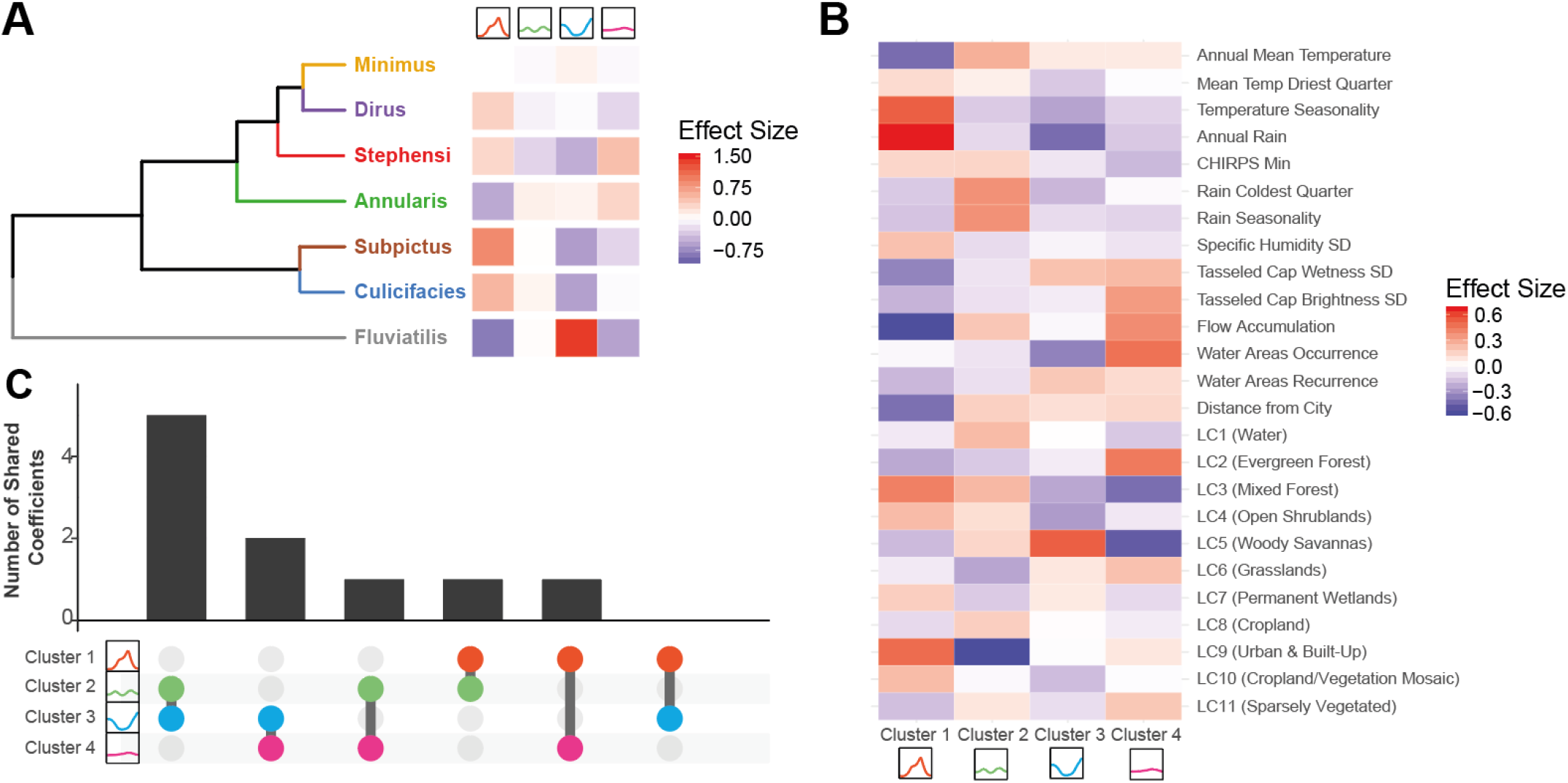
Exploring Drivers of Mosquito Population Dynamics Using Multinomial Logistic Regression. A multinomial logistic regression-based approach using both species complex and a suite of environmental variables was used to explore the factors associated with different mosquito population dynamics. The output of this regression is a single coefficient describing the strength of the association per variable and cluster. **(A)** Hierarchical clustering of the regression results for each species complex, as defined by the set of coefficient values describing the strength of the association between that species complex and the particular cluster. **(B)** The strength of the association between each of the 25 environmental covariates used and the relevant temporal cluster. **(C)** Upset plot summarising the environmental variable coefficients. For each cluster, the 15 environmental covariates with the strongest association were selected and the extent of overlap in this top 15 covariates compared across clusters; x-axis indicates the specific pairwise cluster comparison, y axis the number of shared top 15 covariates between the two clusters.

A number of environmental covariates also demonstrated cluster-specific associations **(Fig.3B)**. Both temperature seasonality and total annual rainfall strongly associated with Cluster 1 (which possessed the dynamics most strongly correlated with rainfall, **Supp Fig.4**). By contrast, perennial dynamics (Cluster 4) strongly associated with the continuous presence of water bodies and negatively associated with both temperature seasonality and rain seasonality. Strong associations with landcover were observed for Cluster 2 (strongly negative for urbanicity) and Cluster 3 (strongly positive for woody savannas). In order to examine the broader patterns of association, we ranked the coefficients for each environmental variable within each cluster according to their magnitude, and selected the 15 with the strongest association in each cluster (positive or negative). The top 15 variables for each cluster were then compared to assess the extent of overlap, revealing that each cluster tended to associate with a unique set of ecological factors **(Fig.3C)**. These mutually exclusive and cluster-specific patterns of association with environmental covariates were similarly borne out across an analysis of the correlation of all coefficients between clusters, which revealed them to be highly negatively correlated **(Supp Fig.5)**.

### Predictive Mapping Highlights the Extensive Variation in Mosquito Dynamics Across India

We next integrated these results with spatial predictions of mosquito species complex presence/absence to produce predictive maps of mosquito population dynamics across India; specifically, to generate estimates of the probability that a given location contains ≥1 mosquito species complex displaying a particular temporal pattern **(Fig.4)**. Our results predict that monsoon peaking dynamics (Cluster 1) are most likely in the North and Northeast **(Fig.4A)**. This contrasts with the predicted spatial distribution of bimodal dynamics (Cluster 2), which are predicted to be more likely across central India and less likely in the Northeast. Dynamics involving peaks during the dry season tracks the predicted spatial distribution of *Anopheles fluviatilis s.l.* closely and are predicted to be most probable across central India **(Fig.4C)** – a similar pattern was observed for spatial predictions of perennial dynamics **(Fig.4D)**. Together these results suggest that spatial variability in both species complex occurrence and environmental factors together generate the complex patterns of mosquito temporal dynamics observed across India.

**Figure 4:**
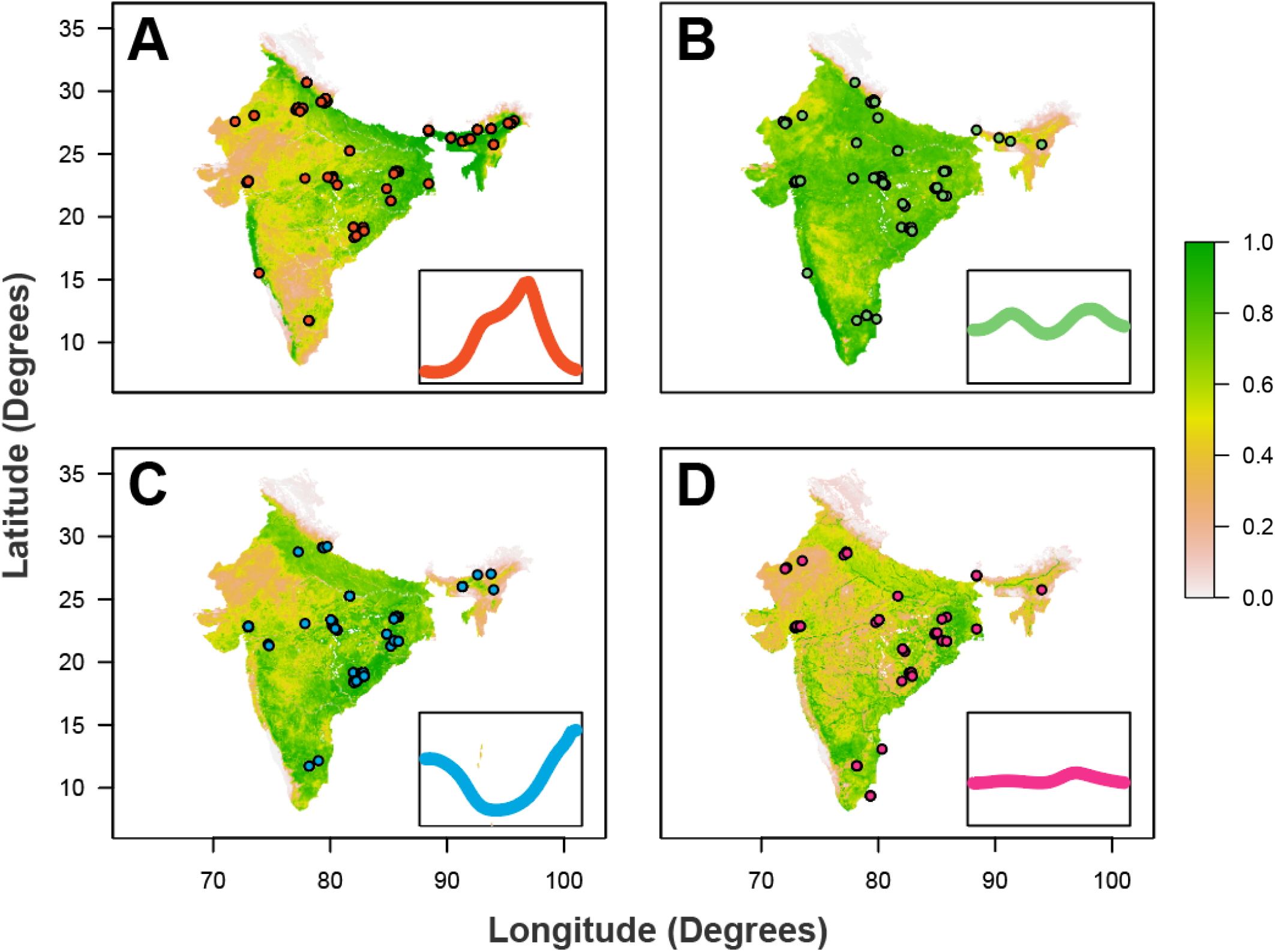
Predictive Maps of Mosquito Population Seasonality Across India. The results of the multinomial logistic regression were integrated with recently generated maps describing the probability of presence/absence for different anopheline species complexes (not shown). Together, these were used to generate estimates of a given area possessing at least one mosquito species complex with a particular temporal profile (as defined by the previously described clusters). **(A)** Results of this analysis for Cluster 1 (the “monsoon peak” cluster) – red dots describe the locations in which a mosquito species complex with a temporal profile assigned to Cluster 1 were found. **(B)** As for A, but for the “bimodal” cluster. **(C)** As for A, but for the “peak in dry season” cluster. **(D)** As for A, but for the “perennial” cluster. In all cases, the map colour describes the probability of a given area containing one or more mosquito species complex displaying that pattern of temporal dynamics. The coloured points indicate locations where a mosquito species complex displaying temporal dynamics belonging to that cluster were empirically observed.

## Discussion

Understanding the temporal dynamics of malaria transmission represents an important input to effective deployment of control interventions. Here we leverage a collection of temporally disaggregated mosquito time-series catch data from across India to explore these dynamics and the comparative role of abiotic and species-specific factors in shaping them. Our results reveal extensive variation in mosquito population dynamics between species complexes and across locations, ranging from highly seasonal and rainfall-concordant dynamics through to perennial and rainfall-discordant dynamics. Analysis of this variation has revealed a complex interplay between biotic (species complex-specific drivers) and abiotic (the broader ecological structure of the environment) factors in shaping these dynamics. Importantly, the comparative importance of these factors depends intimately on the setting and mosquito species complex being considered.

In a manner largely independent of the ecological setting, *Anopheles fluviatilis s.l.* populations typically peaked during the dry season. Whilst previous work has identified these dynamics^38,39^, our work highlights the consistency of this observation across locations, showing that these dynamics are largely restricted to *Anopheles fluviatilis s.l.* and highlight the capacity for the population dynamics of a regionally important malaria vector to significantly depart from local patterns of rainfall. These results align with previous work that has indicated streams and surrounding stagnant water as breeding sites for this species complex^40^ – such breeding sites are typically unsuitable during the monsoon season when flooding occurs but become increasingly suitable as the dry season ensues. By contrast, *Anopheles culicifacies s.l.* displayed a wide array of temporal dynamics depending on the sampling site. These ranged from peaking during the monsoon to bimodal and even perennial behaviour – a finding consistent with documented variation in the species complex*’*s breeding habits^41–43^. However, due to our inability to disaggregate time-series according to sibling species (which frequently show differences in their association with different types of breeding site^44^), the drivers of the observed variation in temporal dynamics for *Anopheles culicifacies s.l.* remains unclear – specifically, whether this diversity is driven by sibling species displaying distinct temporal dynamics or because *Anopheles culicifacies s.l.* temporal dynamics are more plastic and responsive to environmental factors than *Anopheles fluviatilis s.l.* (where the same dynamics were observed irrespective of the broader ecological structure of the surrounding environment).

Our results also support a significant role for the environment in shaping mosquito population dynamics. Perhaps most notably, they highlight limited utility of considering rainfall alone when trying to understand and predict temporal patterns of mosquito abundance. Many of the populations studied here lacked the frequently assumed positive relationship with rainfall and instead displayed patterns of abundance that were only weakly or even negatively correlated. Rainfall is frequently considered a key driver of mosquito population dynamics but the role of temperature (which has a significant influence on many individual mosquito life-history traits^26,45^) in shaping mosquito population dynamics is increasingly being recognised^39^. We identified a significant impact of temperature on population dynamics, with temperature seasonality strongly positively associated with the highly seasonal, monsoon peaking seasonal dynamics (Cluster 1). By contrast, both temperature seasonality and rainfall seasonality were negatively associated with perennial (Cluster 4) dynamics. Together, these results suggest a role for both in shaping annual patterns of mosquito abundance and underscores the importance of considering seasonal fluctuations in temperature, not just rainfall, when trying to understand seasonality in mosquito population dynamics.

The perennial patterns of abundance observed for Cluster 4 were also strongly associated with flow accumulation and water area occurrence (acting as proxies for proximity to rivers and static bodies of water respectively). These factors were negatively associated with all other temporal profiles. This is consistent with reports indicating that static water sources may provide sites available for oviposition and mosquito breeding year round^46,47^ and highlights the importance of the local hydrological environment (which in the cases of large bodies of water is only partially dependent on patterns of rainfall) in shaping mosquito populations and their annual dynamics. We also observed a significant influence of landcover patterns on temporal dynamics. Urbanicity (measured by the two covariates Landcover and Distance to City) was consistently and positively associated with rainfall concordant, monsoon peaking dynamics (Cluster 1) and negatively associated with other temporal profiles. This is possibly due to the diverse array of physical features present in cities (ranging from tyres to wells and overhead tanks) that are able to hold water following rainfall, and which have previously been characterised as breeding sites for a range of mosquito species^48,49^. Overall, these results demonstrate clear structuring of the environmental factors shaping mosquito population dynamics and highlight that unique sets of ecological factors driving each of the different temporal profiles.

It is important to note that factors other than mosquito dynamics are also involved in defining the temporal profile of malaria risk. Whilst an association between the size of mosquito populations and case numbers is well established^50,51^, the nature of this relationship remains less clear. Interactions between malaria endemicity^52^, mosquito abundance^53^ and vector competence^28^ can lead to non-linear dynamics that can be further modified by human behavioural factors such as migration or occupational practices^54^. Due to heterogeneity in mosquito sampling methods and limitations on the extent of entomological data describing relevant malaria metrics such as sporozoite positivity, we were unable to explore many of these factors. Similarly, the lack of disaggregation according to sibling species (which vary markedly in malaria vectorial efficiency) and accompanying epidemiological information (on malaria prevalence or incidence) precludes us from better resolving the comparative contributions of different mosquito species to transmission. This limits our ability to translate temporal patterns of mosquito populations into relevant metrics such as the Entomological Inoculation Rate (EIR). Whilst we mitigate this limitation somewhat by focussing our analyses specifically on dominant vector species-complexes previously established as relevant to malaria transmission in India^55^, it is not necessarily the case that each mosquito species analysed here is equally relevant to malaria transmission. Future work integrating these analyses with those exploring seasonality of case incidence (c.f. Nguyen et al.^31^) would therefore likely prove instructive.

Overall, our work highlights that temporal variation in mosquito populations is driven by a complex interplay of biotic and abiotic factors, with the comparative importance of these depending intimately on the species complex and ecological setting being considered. In doing so, this work underscores the crucial importance of integrating both species composition and ecological structure into our understanding of the temporal profile of malaria risk – a crucial and operationally relevant input for optimising the delivery of malaria control interventions.

## Data Availability

Data will be made open access and fully available upon publication in a peer-reviewed journal.

## Data and Code Availability

All data collaed as part of this study is available in the Supplementary Information and all code used to produce these analyses at https://github.com/cwhittaker1000/anopheleseasonality.

## Acknowledgements and Funding Sources

C.W. is supported by a Medical Research Council Doctoral Training Partnership PhD Studentship. SB & AG both acknowledge grant support from the Bill and Melinda Gates Foundation.

## Author Contributions

CW and SB conceived the study. PW, AG and AK contributed to the design of the study. CW carried out the systematic review and subsequent analyses, with input from SB, AG, AK, PW, MN and additional input, data curation and analysis from MS, SP and CM. CW wrote the manuscript, with PW, MS, SP, CM, DW, MN, PG, AK, AG and SB providing feedback and suggestions during manuscript drafting. All authors approved the final version of the manuscript.

## Methods

### Systematic Review of Indian Entomological Literature

Web of Science and PubMed databases were searched on 17^th^ October 2017 using the keywords “India” AND “Anophel*” to identify references with temporally disaggregated entomological data. We identified 1945 records with 1556 remaining after removing duplicates. Following Title and Abstract screening 281 records were retained for full text evaluation. We included records containing temporally disaggregated adult mosquito catch data with monthly (or finer) temporal resolution spanning at least 12 months that had not been conducted as part of vector control intervention trials, and where sufficient information to geolocate the catch site was provided. 78 references were retained that yielded 117 geolocatable areas across India. These references contained 272 time-series spanning the malaria vectors *Anopheles annularis s.l., culicifacies s.l., dirus s.l., fluviatlis s.l., minimus s.l., stephensi s.l.* and *subpictus s.l.*. See **Supplementary Information** for further details.

### Time-Series Fitting and Interpolation

To smooth the noise in the mosquito catch data we fitted a Gaussian Process model to each of the extracted time-series, using a Negative Binomial likelihood to account for overdispersion in the data:

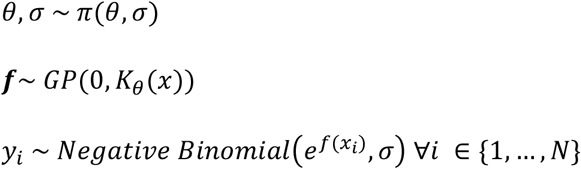

where ***f*** is a distribution of functions from a zero-mean Gaussian Process with covariance function *K*_*θ*_, *f*(x) are function evaluations at times *x, y* are the observed mosquito counts indexed by timepoint *i*, and *σ*and *θ* represent a vector of hyperparameters involved in defining the overdispersion of the Negative Binomial distribution and the functional form of the covariance function respectively. Given that mosquito population dynamics are typically characterised by repeating patterns occurring either seasonally or annually, a periodic kernel function was used to define the covariance between pairs of points, defined as:

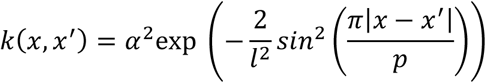

where *p* represents the period over which we would expect points to show similar dynamics (i.e. a period of twelve would imply we expect points separated by 12 months to be most similar), *α* specifies the magnitude of the covariance, and *l* represents a lengthscale parameter further constraining the extent to which two values separated by a given time can co-vary. Weakly informative priors were used although the results were not sensitive to the choice of prior (see **Supp. Fig.4)**. Fitting was undertaken with the probabilistic programming language STAN^29^.

### Time-Series Characterisation and Clustering by Features

Motivated by previous work providing a framework to statistically characterise the empirical structure of time-series data^30^ and work characterising the seasonality of malaria case incidence^31^, we calculated several summary statistics for each smoothed time-series to characterise their temporal properties. These include the Kullback-Liebler divergence (measuring the divergence of the time-series from a uniform distribution), the median of the period (*p*) from the Negative Binomial Gaussian Process fitting (informing the dominant temporal modality present in the data), the proportion of points greater than 1.65x the mean (measuring how peaked the time-series is), the distance of the first peak from January, and then 3 features arising from fitting 1 and 2 component Von-Mises distributions to the smoothed time-series: specifically, the mean of the 1 component Von-Mises distribution, the number of peaks (determined by comparing the quality of fit for 1 and 2 component Von-Mises distributions), and the weight (*ω*), specifying the comparative contributions of each component in the two-component fitting. See **Supplementary Information** for further details. From this we obtain a series of 7 real numbers describing the temporal properties of each time-series. We then applied a Principal Components Analysis to these results to identify a lower-dimensional representation of the structure present in the data amenable to visualisation and implemented k-means clustering to identify clusters of time-series with similar temporal features – i.e. this clustering assigns each smoothed time-series to one cluster.

### Statistical Modelling and Prediction of Seasonal Modality

For each of the 117 study locations we extracted a suite of environmental variables derived from satellite data that together describe the location*’*s ecological structure. These include the BioClimatic variables (a suite of biological relevant covariates defined from monthly rainfall and temperature satellite data^32^), various measures of aridity^33,34^, a number of covariates describing the seasonality and extent of water bodies^35^, landcover^36^ and a number of other variables previously used in defining the global distribution of anopheline vectors^37^. A complete list of the covariates used is **in Supplementary Table 2**. These covariates (25 in total) and a covariate for anopheline species (1 for each time-series indicating which species it belonged to) were used as covariates in a penalised (L2) multinomial logistic regression model predicting the cluster (of time-series with similar temporal properties, assigned based on the results of the k-means clustering) a particular time-series belonged to. Fitting this model yielded regression coefficients describing the strength of association between a species complex/environmental variable and membership of a particular cluster – specifically, 1 coefficient per cluster and predictor, i.e. a total of *k* coefficients per predictor where *k* is the number of clusters. The results of these analyses were then integrated with recently produced maps of vector presence/absence (as part of work conducted with the Humbug Project (http://humbug.ac.uk/), funded through a Google Impact Challenge grant) to generate predictive maps of mosquito population dynamics across India (see **Supplementary Information** for further detail).

