## Supplementary Information for "The Ecological Structure of Mosquito Population Seasonal Dynamics"

#### Contents:


##### Supplementary Information 1: Description of Systematic Review, Data Extraction and Initial Processing

##### Supplementary Information 2: Description of Statistical Methodologies Utilised

##### Supplementary Information 3: Additional Figures and Results

##### References

### Outline of Document

In this supplementary document we outline the methods and data used to explore and analyse the patterns and drivers of *Anopheles* mosquito population dynamics across the Indian sub-continent. In Supplementary Information 1, we present an overview of the systematic search strategy employed, as well as details about the data collated and the initial pre-processing applied to it. In Supplementary Information 2, we detail the statistical methodologies employed to process this extracted data, methodologies whose output forms the basis for the results presented in the main text. Finally, in Supplementary Information 3, we present an array of figures to support the work detailed in the main text.

### Supplementary Information 1: Description of Systematic Review: Data Extraction and Initial Pre-Processing

#### Systematic Review: Search Procedure and Record Screening

Web of Science and PubMed databases were searched on 17th October 2017 using the keywords "India" AND "Anophel*" in order to identify references containing temporally disaggregated entomological data. Our searches identified a total of 1945 records, with 1556 remaining after duplicate removal. References were selected for Inclusion/Exclusion according to the following criteria:

**Inclusion Criteria:**

- Reference contains temporally disaggregated adult mosquito catch data at a temporal resolution of monthly or higher.

**Exclusion Criteria:**

- Mosquito catch data is not temporally disaggregated to a sufficient extent (e.g. catches were done yearly or seasonally rather than monthly).
- Mosquito catch data was collected as part of a trial assessing a vector control intervention (which would perturb the natural dynamics of the vector, rendering the data unrepresentative of the population dynamics in the absence of control).
- Reference only contains information on immature/larval mosquito life cycle stages.
- Reference contained insufficient information to geolocate the area in which the study was conducted.

Following Title and Abstract screening, a total of 281 records were identified, with 78 references retained after Full Text Evaluation. These 78 references contained temporally disaggregated *Anopheles* catch information at monthly resolution (no references were identified which presented higher temporal resolution catch data) for a total of 117 distinct and geolocatable areas across the Indian subcontinent. These form the basis for the results presented in this paper. The next section goes into further detail about extraction and collation of the data associated with each study.

#### Systematic Review: Data Extraction, Collation and Initial Processing

##### Entomological Data Extraction

For each reference, we extracted all relevant entomological catch data detailed. We restricted extraction to 7 major *Anopheles* species known to be relevant to malaria transmission in India (although a number of others exist) and for which multiple catch data time series were available. These were *Anopheles annularis*[1], *Anopheles culicifacies*[2,3], *Anopheles dirus*[4,5], *Anopheles fluviatilis*[6,7], *Anopheles minimus*[8,9], *Anopheles stephensi*[10] and *Anopheles subpictus*[3,11]. Where data were presented in the form of a table, data was copied directly from the table. Where graphs only were presented, estimates of the data were extracted using DataThief™ software. This yielded a total of 305 time series of monthly mosquito catch data, ranging in length from 5 – 46 months. We restricted subsequent analyses to time series that spanned a year (12 timepoints, monthly) or longer, a total of 272 time series. This yielded the following number of time series for each of the species considered (Supplementary Table 1):

**Supplementary Table 1: Summary of the Number of Time Series Extracted, Disaggregated By Species**

| | *A.annularis* | *A.culicifacies* | *A.dirus* | *A.fluviatilis* | *A.minimus* | *A.stephensi* | *A.subpictus* |
|---|---|---|---|---|---|---|---|
| **# Time Series** | 39 | 85 | 11 | 60 | 12 | 27 | 38 |

As the primary focus of this research was to explore annual and seasonal patterns of mosquito population dynamics, as well as the fact that variations in time series length are a factor known to affect their statistical properties[12] (and which would therefore impact the comparability of the time series gathered and analysed here), all time series were standardised to be 12 months in length. For time series containing more than 12 time points (i.e. time series that spanned longer than a single year), we averaged the recorded catches for a given month. Where the study has been initiated in a month other than January, and concluded in a month other than December, the recorded counts were rearranged to yield a complete time series running from January to December. The studies analysed here employed a wide array of different sampling methodologies including Indoor and Outdoor Resting Collections, Human Landing Catches, Spray Catches and Trap Catches amongst others. Results were typically, though not always, presented in the form of some sampling-effort standardised measure such as Man Hour Density (MHD). As such, though reflective of mosquito population dynamics, these measures do not represent the overall number of mosquitoes caught. To this end, where information on sampling effort (number of hours spent sampling, number of households/cattlesheds searched, number of human baits, number of traps set etc) was present, we used this information to convert MHD back to the raw counts. In the small number of instances where there was variable sampling effort across the time series (which would bias the conversion away from the true underlying population dynamics), we conservatively used the lowest sampling effort recorded across the time series in the conversion. Together, this allowed us to produce an estimate of the number of mosquitoes sampled (a raw count, based on equal sampling effort across the time series). See Supplementary Data: Temporal Information, sheet Raw Catch Data for more information about the transformations applied to each of the time series, as well as the unprocessed catch data extracted from each of the references.

**Environmental Covariate Assembly**

The environmental covariates (i.e. the independent variables that, along with species, are used to predict the different seasonal patterns) used in this research consist of raster layers spanning all of India at a 2.5 arc-minute (~ 5km by 5km) spatial resolution. The covariates utilised here were initially selected from based a set of 66 covariates derived from:

- Covariates previously used in other *Anopheles* mosquito mapping efforts[13], as well as in other mapping efforts looking at the spatial distribution of the malaria parasite, *Plasmodium falciparum*[18].
- Consideration of some of the possible drivers of seasonal dynamics (primarily hydrological considerations surrounding the seasonality and availability of aquatic breeding sources, and how this might interact with environmental composition[14,15] and species specific breeding preferences[16,17] to structure population dynamics). From these considerations, a number of other raster layers were included that together describe further the underlying hydrological environment.

The majority of these covariates are derived from high temporal resolution satellite images that were initially gap-filled[19] to eliminate missing data that typically arises from cloud cover. These images were then aggregated and summarised to produce a suite of synoptic environmental covariates for prediction. From these 66 covariates (a number of which are highly correlated with one another), a reduced subset of 25 covariates were selected. These were selected in the following way. Firstly covariates were grouped into one of five categories based on the ecological features they were describing. These categories were Temperature, Rainfall, Aridity, Hydrological and Landcover. A subset of covariates were then selected in each category in order to minimise the correlation between covariates (based on correlation matrices of Spearman correlation coefficients) whilst also retaining measures of important quantities such as the mean, the dispersion etc for a given category. Based on this, the final covariates included in each category were the following:

- **Temperature:** Annual Mean Temperature, Temperature Seasonality & Mean Temperature in the Driest Quarter (3 covariates).
- **Rain:** Annual Rain, Rain Seasonality, CHIRPS Minimum and Rain in the Coldest Quarter (4 covariates).
- **Aridity:** Specific Humidity Standard Deviation, Tasseled Cap Wetness Standard Deviation, Tasseled Cap Brightness Standard Deviation (3 covariates).
- **Hydrological:** Water Areas Occurrence, Water Areas Recurrence and Flow Accumulation (3 covariates).
- **Landcover:** Dominant Landcover and City Accessibility (2 covariates).

A total of 25 covariates (Dominant Landcover consists of 11 classes of landcover type). This reduction reduced the extent of multicollinearity in the covariates, reduced the scope for model overfitting, and had minimal impact on the predictive power (a correct classification rate of 60% for the model including all 66 covariates, which reduced to 58% when using the reduced subset of 25 covariates).

**Study Geolocation and Environmental Covariate Extraction**

Geolocation of study areas was possible to a varying degree depending on the information available within the paper (and related literature). When villages names or the details of the administrative unit a study was carried out in were provided in the paper text, geolocation was carried out utilising a wide array of resources containing spatially explicit information on the location of Indian settlements and administrative units. These were Google Maps/Google Earth, Etrace, OneFiveNine, Veethi, Wikimapia, VillageInfo, MapsOfIndia, Geonames and AlipurduarTourism. Additionally, a number of the references identified in our review had previously been utilised as part of the Malaria Atlas Project (MAP) Presence/Absence mapping work and so had previously been geolocated[20]. In these instances, the MAP location estimate was used. The precision of study location estimates varied greatly (due to the extent of spatial detailed provided in the paper e.g. village vs district as well as the identifiability of villages/administrative units) – this uncertainty is explicitly incorporated into our analyses, with raster covariates extracted over the full area the study is believed to have been carried out in, and then the average of those raster values used. In addition to the environmental covariates detailed above, for each of the 117 geolocated study locations, daily rainfall estimates spanning the sampling period were also collated. These data were taken from "The Climate Hazards Group Infrared Precipitation With Stations" (CHIRPS) dataset[21] and were subsequently aggregated up to the same temporal resolution as the mosquito catch data. Data from the CHIRPS dataset is only available from the year 1981, and so for locations where the sampling date predated this, daily rainfall data was extracted for the year 1981, and assumed to be representative of past rainfall. See Supplementary Data Overall Temporal Information, Location & Spatial Information for more information about the specific resources used to geolocate each individual study location.

**Maps of Vector Presence/Absence**

Extensive work has previously been undertaken mapping the distributions of key *Anopheline* vectors across Africa, the Middle East and Europe[22], the Americas[23] and the Asia and Pacific region[20]. These maps describe the probability of occurrence at a 5km by 5km resolution for many of the dominant vector species involved in malaria transmission. Here, we utilise updated versions of these maps that include presences up to the year 2016 as part of work conducted with the Humbug Project (http://humbug.ac.uk/), funded through a recent Google Impact Challenge grant. These maps describe the probability of occurrence for the species *An. annularis, An. culicifacies, An. dirus, An. fluviatilis, An. minimus, An. stephensi* and *An. subpictus*. These estimates of occurrence probability were then integrated with a multinomial logistic regression model of dynamics to generate estimates of the probability of a given location containing a particular temporal pattern/profile (see section **Penalised**

**Multinomial Logistic Regression Modelling, Evaluation of Model Accuracy and Predictive Modelling** below for further technical details).

**Supplementary Table 2: Environmental Covariates Explored in the Variable Selection Process and Utilised in Modelling and Prediction of Seasonal Population Dynamics**

| # | Variable | Temporal Resolution | Source |
|---|---|---|---|
| 1 | Annual Mean Temperature | Annual Average, 1970 - 2000 | https://www.worldclim.org/bioclim |
| 2 | Mean Diurnal Range | Annual Average, 1970 - 2000 | https://www.worldclim.org/bioclim |
| 3 | Isothermality | Annual Average, 1970 - 2000 | https://www.worldclim.org/bioclim |
| 4 | Temperature Seasonality | Annual Average, 1970 - 2000 | https://www.worldclim.org/bioclim |
| 5 | Max Temperature of Warmest Month | Annual Average, 1970 - 2000 | https://www.worldclim.org/bioclim |
| 6 | Min Temperature of Coldest Month | Annual Average, 1970 - 2000 | https://www.worldclim.org/bioclim |
| 7 | Temperature Annual Range | Annual Average, 1970 - 2000 | https://www.worldclim.org/bioclim |
| 8 | Mean Temperature of Wettest Quarter | Annual Average, 1970 - 2000 | https://www.worldclim.org/bioclim |
| 9 | Mean Temperature of Driest Quarter | Annual Average, 1970 - 2000 | https://www.worldclim.org/bioclim |
| 10 | Mean Temperature of Warmest Quarter | Annual Average, 1970 - 2000 | https://www.worldclim.org/bioclim |
| 11 | Mean Temperature of Coldest Quarter | Annual Average, 1970 - 2000 | https://www.worldclim.org/bioclim |
| 12 | Annual Precipitation | Annual Average, 1970 - 2000 | https://www.worldclim.org/bioclim |
| 13 | Precipitation of Wettest Month | Annual Average, 1970 - 2000 | https://www.worldclim.org/bioclim |
| 14 | Precipitation of Driest Month | Annual Average, 1970 - 2000 | https://www.worldclim.org/bioclim |
| 15 | Precipitation Seasonality | Annual Average, 1970 - 2000 | https://www.worldclim.org/bioclim |
| 16 | Precipitation of Wettest Quarter | Annual Average, 1970 - 2000 | https://www.worldclim.org/bioclim |
| 17 | Precipitation of Driest Quarter | Annual Average, 1970 - 2000 | https://www.worldclim.org/bioclim |
| 18 | Precipitation of Warmest Quarter | Annual Average, 1970 - 2000 | https://www.worldclim.org/bioclim |
| 19 | Precipitation of Coldest Quarter | Annual Average, 1970 - 2000 | https://www.worldclim.org/bioclim |
| 20 | Potential Evapotranspiration | Annual Average, 1950 - 2000 | https://cgiarcsi.community/data/global-aridity-and-pet-database/ |
| 21 | Global Aridity Index | Annual Average, 1950 - 2000 | https://cgiarcsi.community/data/global-aridity-and-pet-database/ |
| 22 | Population Density | 2010 | http://www.worldpop.org.uk |

| | | | |
|---|---|---|---|
| 23 | Day Land Surface Temperature Mean | Annual Average 2001 – 2015 | https://developers.google.com/earth-engine/datasets/catalog/Oxford_MAP_LST_Day_5km_Monthly |
| 24 | Day Land Surface Temperature SD | Annual Average 2001 – 2015 | https://developers.google.com/earth-engine/datasets/catalog/Oxford_MAP_LST_Day_5km_Monthly |
| 25 | Night Land Surface Temperature Mean | Annual Average 2001 – 2015 | https://developers.google.com/earth-engine/datasets/catalog/Oxford_MAP_LST_Night_5km_Monthly |
| 26 | Night Land Surface Temperature SD | Annual Average 2001 – 2015 | https://developers.google.com/earth-engine/datasets/catalog/Oxford_MAP_LST_Day_5km_Monthly |
| 27 | Tasselled Cap Wetness Mean | Annual Average 2001 – 2012 | https://developers.google.com/earth-engine/datasets/catalog/Oxford_MAP_TCW_5km_Monthly |
| 28 | Tasselled Cap Wetness SD | Annual Average 2001 – 2012 | https://developers.google.com/earth-engine/datasets/catalog/Oxford_MAP_TCW_5km_Monthly |
| 29 | Tasselled Cap Brightness Mean | Annual Average 2001 – 2012 | https://developers.google.com/earth-engine/datasets/catalog/Oxford_MAP_TCB_5km_Monthly |
| 30 | Tasselled Cap Brightness SD | Annual Average 2001 – 2012 | https://developers.google.com/earth-engine/datasets/catalog/Oxford_MAP_TCB_5km_Monthly |
| 31 | Elevation | NA | https://developers.google.com/earth-engine/datasets/catalog/USGS_SRTMGL1_003 |
| 32 | Flow Accumulation | NA | https://developers.google.com/earth-engine/datasets/catalog/WWF_HydroSHEDS_15ACC |
| 33 | Specific Humidity Mean | Annual Average 1948 - 2010 | https://developers.google.com/earth-engine/datasets/catalog/NASA_GLDAS_V20_NOAH_G025_T3H |
| 34 | Specific Humidity SD | Annual Average 1948 – 2010 | https://developers.google.com/earth-engine/datasets/catalog/NASA_GLDAS_V20_NOAH_G025_T3H |
| 35 | Enhanced Vegetation Index | Annual Average 2001 – 2015 | https://developers.google.com/earth-engine/datasets/catalog/Oxford_MAP_EVI_5km_Monthly |
| 36 | Landcover | Annual Dominant Value 2001 – 2012 | https://developers.google.com/earth-engine/datasets/catalog/Oxford_MAP_IGBP_Fractional_Landcover_5km_Annual |
| 37 | Water Body Maximum Extent | 1984 – 2015 | https://developers.google.com/earth-engine/datasets/catalog/JRC_GSW1_0_GlobalSurfaceWater |
| 38 | Water Body Seasonality | Average 1984 – 2015 | https://developers.google.com/earth-engine/datasets/catalog/JRC_GSW1_0_GlobalSurfaceWater |
| 39 | Water Areas Occurrence | Average 1984 – 2015 | https://developers.google.com/earth-engine/datasets/catalog/JRC_GSW1_0_GlobalSurfaceWater |
| 40 | Water Areas Recurrence | Average 1984 – 2015 | https://developers.google.com/earth-engine/datasets/catalog/JRC_GSW1_0_GlobalSurfaceWater |

| | | | |
|---|---|---|---|
| 41 | Digital Chart of the World - Distance to Nearest Water Body | Average 1984 – 2015 | Generated manually using the information on Water Body location available from Digital Chart of the World: |
| 42 | World Wildlife Fund – Distance to Nearest Water Body | Average 1984 – 2015 | Generated manually using the information on Water Body location available from the World Wildlife Fund: |
| 43 | Distance from City | 2015 | https://www.nature.com/articles/nature25181 |
| 44 | CHIRPS Maximum | Derived from CHIRPS rainfall data and the time-period it spans | https://www.nature.com/articles/sdata201566 |
| 45 | CHIRPS Minimum | Derived from CHIRPS rainfall data and the time-period it spans | https://www.nature.com/articles/sdata201566 |
| 46 | CHIRPS Mean | Derived from CHIRPS rainfall data and the time-period it spans | https://www.nature.com/articles/sdata201566 |
| 47 | WC A0 | Component of Fourier transform of rainfall data – amplitude | Malaria Atlas Project |
| 48 | WC A1 | Component of Fourier transform of rainfall data – amplitude | Malaria Atlas Project |
| 49 | WC A2 | Component of Fourier transform of rainfall data – amplitude | Malaria Atlas Project |
| 50 | WC A3 | Component of Fourier transform of rainfall data – amplitude | Malaria Atlas Project |
| 51 | WC P0 | Component of Fourier transform of rainfall data – frequency | Malaria Atlas Project |
| 52 | WC P1 | Component of Fourier transform of rainfall data – frequency | Malaria Atlas Project |
| 53 | WC P2 | Component of Fourier transform of rainfall data – frequency | Malaria Atlas Project |
| 54 | WC P3 | Component of Fourier transform of rainfall data – frequency | Malaria Atlas Project |
| 55 | Urban Footprint | Describing the extent of urbanicity in a given area | Malaria Atlas Project |
| 56 | Irrigated Areas | Describing the extent of irrigation in a given area | Malaria Atlas Project |

**Note:** There are 66 covariates total here, as Landcover contains 11 distinct covariates (each describing the proportion of cover attributable to a particular landcover class in a given area).

**Note:** All WorldClim data is from Version 2 of the datasets.

#### Supplementary Information 2: Description of Statistical Methodologies

##### Negative Binomial Gaussian Process – Fitting and Inference:

We use a highly flexible stochastic process model, known as a Gaussian Process, to temporally interpolate between the monthly catch datapoints and integrate over uncertainties in the estimates of mosquito abundance (a product of both the catch methodology as well as generic random variation in the sampling of the mosquito population) spanning the entire year. Gaussian processes specify a distribution over functions such that any finite set of function values $f(x_1), f(x_2), \ldots f(x_N)$ have a joint Gaussian distribution[24]. The Gaussian process is entirely specified by its mean function, defined as:

$$E[f(x)] = \mu(x)$$

and by its covariance function:

$$Cov[f(x), f(x')] = k(x, x')$$

also known as the kernel. This kernel is a positive-definite function of two inputs, $x$ and $x'$ that defines the covariance between any two points (and by extension the covariance matrix of our Gaussian Process when all pairwise combinations of points are considered). In doing so, the kernel encodes prior information about the extent to which we would expect two objects ($x$ and $x'$ in this instance) to be similar. A wide array of kernels exist that specify an equally wide array of similarity structures, such as the squared exponential (where similarity varies with the Euclidean distance separating $x$ and $x'$) and the linear kernel (which allows the relationship governing similarity to vary with not just the relative position of two inputs, i.e. $x - x'$, but with their absolute position, a property that makes this kernel "non-stationary"). Given the strong seasonality known to be present in mosquito catch time series and from the empirically observed patterns of abundance observed when examining the raw time series **(Supplementary Figure 1)**, we selected a Periodic Kernel. This kernel defines similarity based on the distance between $x$ and $x'$ compared to some period $p$ and so is able to accommodate patterns that broadly repeat themselves over time (such as seasonal or annual peaks in mosquito abundance).

$$k(x, x') = \alpha^2 \exp\left(-\frac{2}{l^2} sin^2\left(\frac{\pi|x - x'|}{p}\right)\right)$$

Where the $p$ represents the period, $\alpha$ specifies the magnitude of the covariance given a certain period, and $l$ represents a lengthscale parameter further constraining the extent to which two values separated by a given distance can co-vary with one another.

Bayesian inference and fitting of Gaussian Processes typically utilises the following hierarchical formulation:

$$\theta \sim \pi(\theta)$$

$$f \sim GP(0, K_\theta(x))$$

$$y_i \sim MVN(f(x_i), \sigma^2)\ \forall i\ \in \{1, \ldots, N\}$$

where $\theta$ represents a vector of hyperparameters involved in defining the kernel's properties, $f$ is a distribution of functions from a zero-mean Gaussian Process with covariance function $K_\theta$, $f$(x) are function evaluations at times $x$, and $y$ our observed counts. However, mosquito catch data is rarely normally distributed (leaving aside limit theorems) and frequently displays high levels of overdispersion[25], a common property of biological systems generally, but made more acute by the fact that for a number of the time series, the monthly catches reported represented the summed total of multiple catches made throughout the period (but that were not presented in the paper, where only monthly totals were presented); this process of summation also introduces overdispersion. Motivated by this, we adapted the above framework to accommodate a Negative Binomial likelihood, leading to the following inferential framework:

$$\theta \sim \pi(\theta)$$

$$f \sim GP(0, K_\theta(x))$$

$$y_i \sim Negative\ Binomial\left(e^{f(x_i)}, \sigma\right) \forall i \in \{1, \ldots, N\}$$

where the exponential function $e^x$ is used to reflect the fact that we use a log link between the observed counts and the underlying latent process reflecting the population dynamics, and $\sigma$ represents the overdispersion parameter of the Negative Binomial distribution.

**Prior Probability Specification**

Prior distributions for the estimated parameters were defined as follows:

$$l \sim Normal(2, 1^2)$$

$$\alpha \sim Half - Normal(0, \sqrt{SD\ (y)})$$

$$p \sim Normal(12, 4^2)$$

$$\sigma \sim Half - Normal(0, 8^2)$$

Weakly informative priors were set on the scaling factor $\alpha$, the period, $p$, and the overdispersion parameter, $\sigma$. The period prior was centred around 12 (a value which would represent annual variation) to reflect the fact that the majority of observed variation in mosquito abundance recorded has typically been observed to cycle annually due to annual variation in key ecological factors such as rainfall and ambient temperature **(**a phenomenon supported by the results in **Supplementary Figure 1)**. A wide standard deviation was used however in to allow the model to identify and accommodate instances of bimodality or periods operating across timescales longer than a year, although important to note is that the lower and upper bounds for the period were set to 4 and 18 months respectively, to avoid identifiability issues arising from the lack of data at temporal resolutions substantially below and above these bounds. A similarly wide prior was set over the overdispersion parameter $\sigma$ and the scaling factor $\alpha$. An informative prior was set for the lengthscale $l$, although the use of less informative priors, either for the lengthscale or for the period, did not significantly alter conclusions arising from the analysis (see **Supplementary Figure 3**), highlighting the robustness of the results presented in the main text.

**Model Fitting and Parameter Inference**

This Negative Binomial Gaussian Process were fitted using STAN, a probabilistic programming language for statistical inference written in C++ that employs gradient-based Markov Chain Monte Carlo algorithms (the No-U-Turn sampler, a variant of Hamiltionian Monte Carlo) for Bayesian inference[26]. The model specified above was implemented in R using the rStan package[27]. For each time series, 4 chains of 5,000 iterations were run for purposes of model fitting and parameter inference. Half of each chain's iterations were discarded as burn-in/the adaptive phase of the sampling, leaving a total of 10,000 iterations available for inference. Measures of MCMC convergence such as the Gelman-Rubin statistic were monitored in all cases and were all consistently < 1.02, indicating stability of the chains and probable convergence to the underlying true posterior distribution.

**Fitted Time Series Normalisation and Von Mises Distribution Fitting:**

Following this fitting process, and to establish comparability across the time series (which varied substantially in the absolute count numbers recorded and used a wide and highly heterogeneous array of different sampling methods), we normalised each time series in the following way:

$$p_i = \frac{y_i}{\sum y_i}$$

where $p_i$ is the normalised count for timepoint $i$ and $y_i$ is the un-normalised count for timepoint $i$ as predicted from the Negative Binomial GP fitting described in the previous section.

To further characterise the periodic properties of these time series, we fit a Von Mises distribution, which is a continuous probability distribution on the circle with range from 0 to $2\pi$. Broadly, it can be regarded as the circular analogue of the normal distribution on the line, with the probability density function for the angle $x$ given by:

$$f(x|\mu, \kappa) = \frac{e^{\kappa \cos(x-\mu)}}{2\pi I_0(\kappa)}$$

where $I_0(\kappa)$ is the modified Bessel function of order 0, the parameter $\mu$ is a measure of location (analogous to the mean of the normal distribution, describing where on the circle the distribution is clustered around) and $\kappa$ describes the concentration of density around $\mu$ (and thus its inverse is a measure of dispersion, analogous to $\sigma^2$ for the normal distribution.

We fit two sets of Von Mises densities to the normalised time series, the first containing a single component, specified as:

$$f(x|\mu_1, \kappa_1) = f_1(x|\mu_1, \kappa_1)$$

And the other possessing two components (sometimes called a mixture), formulated as:

$$f(x|\mu_1, \kappa_1, \mu_2, \kappa_2, w) = \omega f_1(x|\mu_1, \kappa_1) + (1-\omega) f_2(x|\mu_2, \kappa_2)$$

where $x$ in both instances represents the normalised monthly count formulated as a random variable on the circle, i.e. by defining $x = \frac{2\pi p_i}{12}$ . Fitting was carried out in R using the *optim* function and with the sum of squares as the loss function. The outputs arising from this fitting – the comparative suitability of the one and two component distributions, as well as the values of $\omega$, $\mu$ and $\kappa$, were then explored to further characterise the temporal properties of the data.

**Time Series Characterisation and Analysis:**

Following this, we sought to characterise the properties and features of the smoothed time series. To do this, we applied a series of mathematical operations to the time series, taking inspiration from recent work carried out exploring the empirical structure of time series arising from a disparate array of different fields[12]. Doing so facilitates comparison of features between time series, and identification of time series sharing similar statistical properties. The operations used were the following:

1. **Kullback-Leibler Divergence:** Also known as the relative entropy, the Kullback-Liebler divergence represents a measure of how different one probability distribution is from a second probability distribution (where a value of 0 indicates that the two distributions are identical). It is specified in the following manner:

$$E_i = p_i log_2\left(\frac{p_i}{q_i}\right)$$

$$E = \sum_{i=1}^{12} p_i log_2\left(\frac{p_i}{q_i}\right)$$

where $p_i$ is the average value of the normalised time series for month $i$, and $q_i$ = 1/12 for $i = 1, \ldots, 12$. This operation therefore measures the deviation of a normalised time series from a uniform distribution, in doing so, informing about the extent to which a seasonal peak (or peaks) is present in the time series.

2. **Periodic Kernel Median:** Fitting the Negative Binomial Gaussian Process with a periodic kernel allowed inference of the period, $p$, providing us with an estimate of the frequency of repeating patterns in the monthly abundance of mosquitoes. An estimate of $p$ was calculated for each fitted time series, with inference based on the 10,000 MCMC iterations described in further detail in the section **Supplementary Information Model Fitting and Parameter Inference.** The median value of $p$ across these 10,000 iterations was used here.
3. **Proportion of Points Greater Than 1.65x the Mean:** For each fitted, normalised time series, the proportion of points greater than 1.65x the mean of the time series was calculated. This informs about the extent to which the data is peaked, as well as the width of the peak.
4. **Peak Distance from January:** For each fitted, normalised time series, the maximum recorded value was noted and the distance of this value from January was calculated.
5. **Number of Peaks:** Estimates of the parameters governing the fitted two component Von Mises distribution were used to infer the number of peaks in each time series (detailed further in the section **Supplementary Information Von Mises Distribution Fitting**). Specifically, a time series was deemed to possess one peak if the value of the Von Mises component weighting was either < 0.3 or > 0.7 and the difference in means was < $\frac{2\pi}{3}$ or > $\frac{4\pi}{3}$ , indicating that the majority of the density could be attributed to one of the two components, and that the two means identified during the fitting were temporally close to one another. Otherwise, a time series was judged to possess two peaks.
6. **Von Mises 1 Component Mean:** This operation is based on the number of peaks inferred from fitting of 1 and 2 component Von Mises distributions to the Negative Binomial GP fitted, normalised time series. If a 1 component Von Mises distribution was preferred, then the Von Mises mean corresponding to the maximum likelihood predicted value was used. If the 2 component Von Mises distribution was preferred, the value for this operation for that particular time series is set to -5.
7. **Von Mises Two Component Weight:** Estimates of the weight parameter governing the two component Von Mises distribution were also used to infer the bimodality of the time series. The weight specifies the proportion of each component that is used to fit the time series and thus a very high (or very low weight) indicates the dominance of a single component and the comparatively small contribution of the other.

**Principal Components Analysis and Clustering:**

PCA is a statistical procedure that utilises an orthogonal transformation to convert a set of correlated variables (in this case the outputs of the 7 mathematical operations described above for each of the time series) into a set of linearly uncorrelated variables (known as the “principal components”). In doing so, this allows us to summarise this set of variables with a smaller number of representative variables that together explain the majority of the variability in the variables. Reducing the dimensionality of the dataset in this way facilitates visualisation of time series properties (as defined by the mathematical operations) as well as clustering of the time series into groups which share similar properties (clustering algorithms typically perform poorly in high dimensional settings, necessitating the use of PCA as described here, see Supplementary Figure 6 for the variation explained as a function of PCA dimensions). Clustering was then undertaken using the k-means clustering algorithm.

**Penalised Multinomial Logistic Regression Modelling, Evaluation of Model Accuracy and Predictive Modelling:**

Prediction of cluster membership was carried out using a multinomial logistic regression model. Multinomial logistic regression generalises logistic regression (which predicts a binary outcome) to instances with >2 possible outcomes and predicts the probabilities of all possible outcomes of a categorically distributed dependent variable (in this instance, the 4 clusters representing distinct temporal patterns) given a set of independent variables (in this instance, the species each time series belongs to and a suite of environmental covariates as defined in **Supplementary Table 2**). Whereas logistic regression frameworks typically have a single coefficient per covariate (which describes the influence of that particular covariate on the outcome being 1 rather than 0), within a multinomial logistic framework, each category being predicted (each of the 4 clusters of temporal patterns in this instance) has a coefficient per covariate. Thus, a given covariate e.g. Isothermality will have 4 coefficients associated with it, with each of these 4 coefficients specifying the association between Isothermality and membership of Clusters 1, 2, 3 and 4 respectively. We employed an $\ell 2$ (ridge) penalty on all coefficients in order to manage and reduce issues surrounding overfitting: this regularised multinomial logistic regression model was then fitted within a Bayesian framework and implemented in STAN. Following fitting, the mean coefficient values were used to generate estimates of the probability that a given time series belongs to each of the 4 clusters. Time series were assigned to the cluster with the highest cluster probability and the misclassification rate computed based on the proportion of time series whose cluster membership was correctly predicted.

This model was then used to predict seasonal profiles across the Indian subcontinent. Specifically, it integrated recently generated maps of vector presence/absence for each of the vector species considered here to produce estimates of the probability of locations containing at least one mosquito species displaying a particular temporal pattern (1 of the 4 temporal patterns associated with the clusters). For each location, we individually calculated for each of the seven species the probability that the species was present (using the recently generated maps of vector presence/absence), and the probability that the species would display a particular temporal pattern, conditional on presence. Specifically, for each temporal profile $TP$, the probability of a vector species $j$ being present and displaying that particular temporal profile was calculated as follows:

$$p(TP_j) = p(TP_j \mid VP_j)\, p(VP_j)$$

where $p(VP_j)$ describes the probability of the vector occurring in that location (taken from the Humbug vector occurrence probability maps) and $p(TP_j \mid VP_j)$ describes the probability of that vector species displaying that temporal profile conditional on its occurrence in that location. Then, treating each of these 7 probabilities as binomial random variables, the probability of a location containing at least one mosquito species displaying this particular temporal pattern is equal the product of 1 – the probability of temporal profile absence for each of the 7 species.

#### Supplementary Information 3: Additional Figures and Results

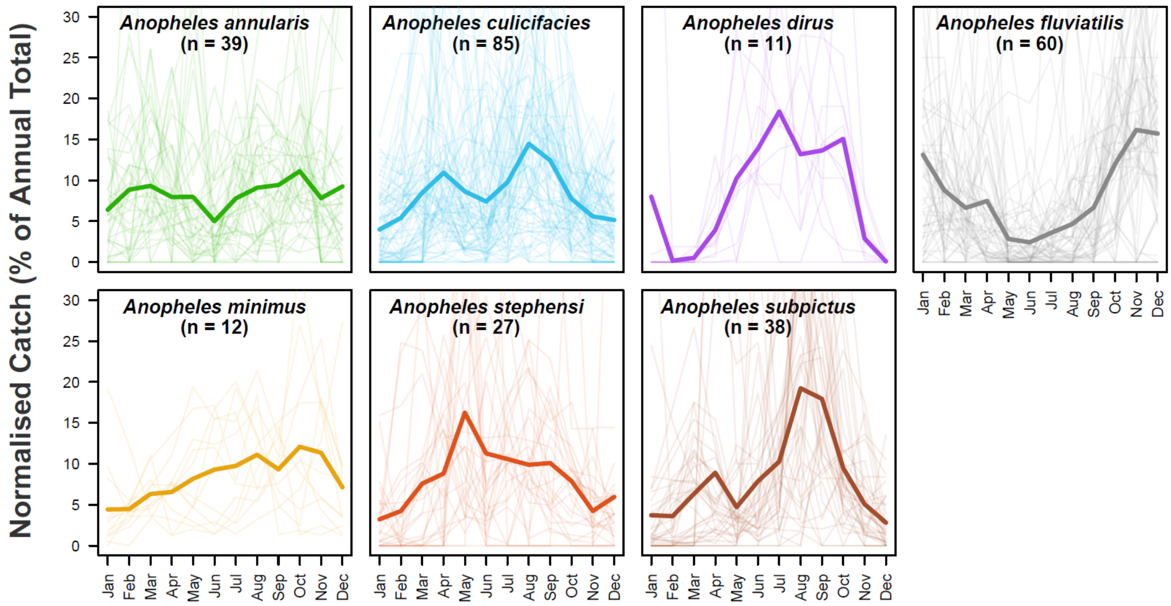


**Supplementary Figure 1: The Raw Mosquito Data Extracted During the Systematic Review Process.** Through a systematic review, a total of 272 time series containing species-specific, monthly disaggregated mosquito catch data spanning at least 12 months were identified and extracted. Together, these time series span 118 locations across India and 7 major *Anopheline* species known to be involved in the transmission of malaria. For each panel presented here, pale lines represent a single normalised time series for that particular species, and the brighter line is the mean of all the time series belonging to that species, evaluated at that particular timepoint.

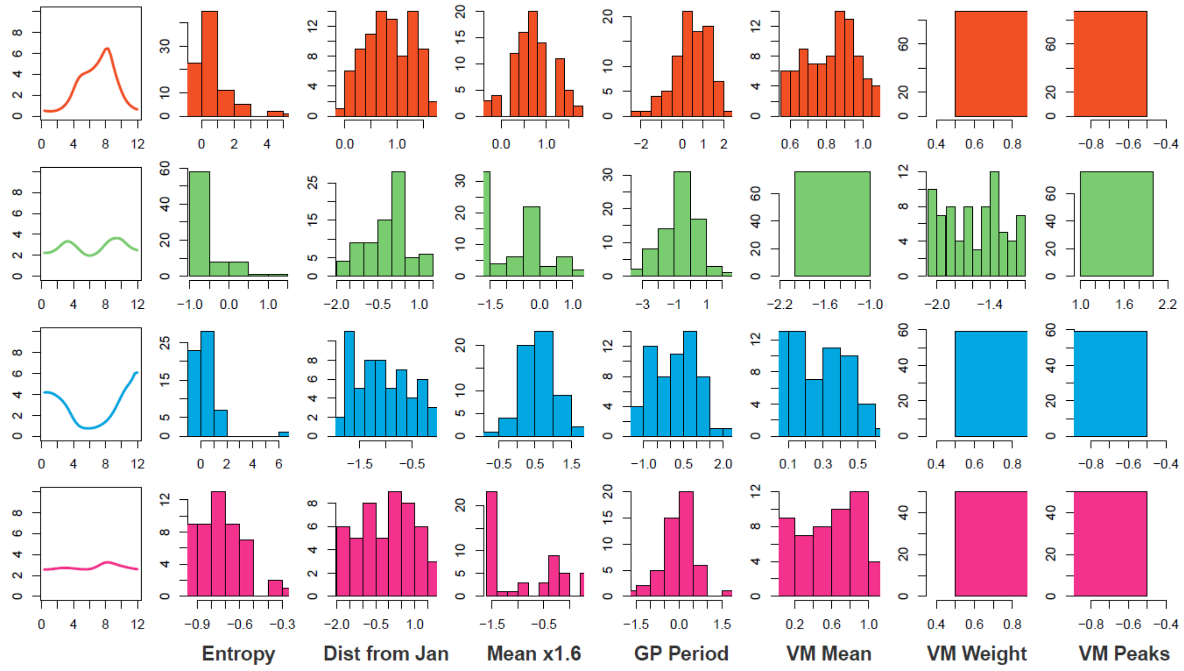


**Supplementary Figure 2: Temporal Cluster Properties.** A series of mathematical operations were applied to the fitted time series in order to further characterise and explore their temporal properties. The results of this characterisation were then clustered using the k-means algorithm. For each cluster, the mean temporal profile is displayed, as well as the underlying distribution of each temporal property is displayed, namely the **Entropy**, the **Distance of the Highest Peak from January**, the **Proportion of Points > 1.6x the Mean,** the **Period of the Fitted Gaussian Process Kernel**, the **Mean of the Fitted Von Mises Distribution**, the **Weights of the Two Von Mises Components** and the **Optimal Number of Von Mises Components.** For further information on each of these operations, see Supplementary Information: Time Series Characterisation and Analysis.

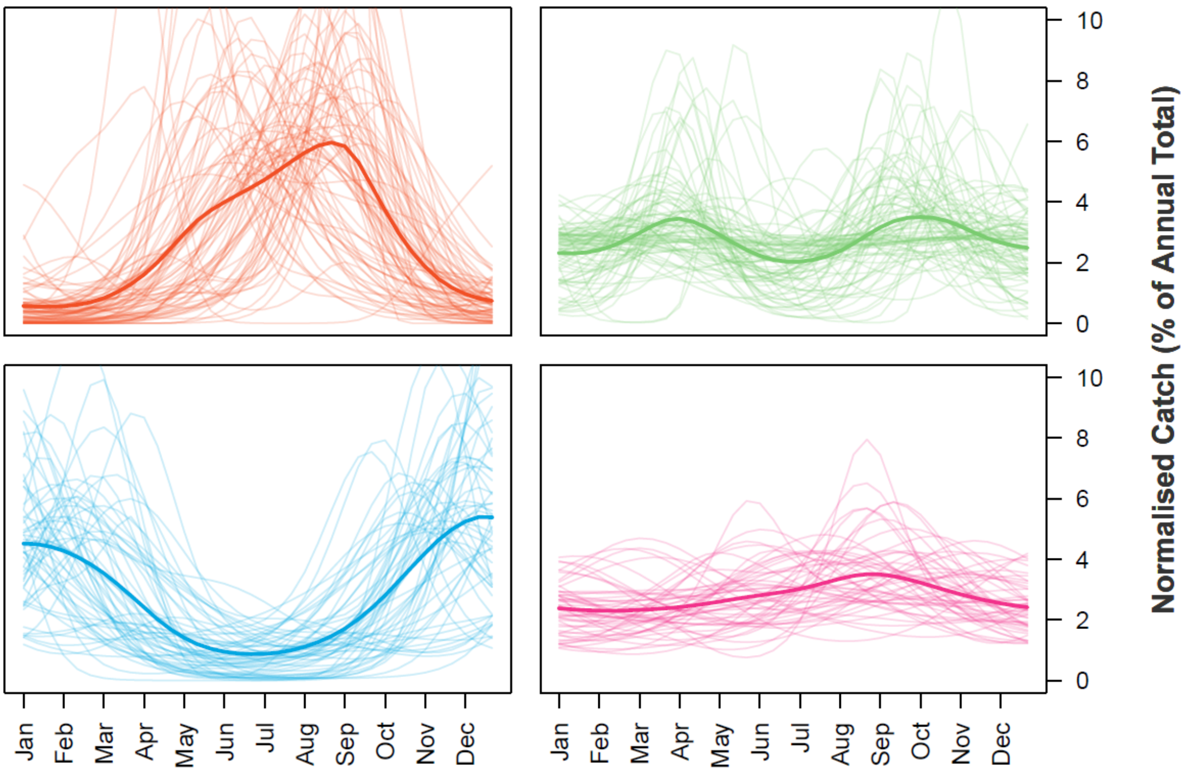


| | Uninformative Prior | | | |
|---|---|---|---|---|
| **Informative Prior** | **Cluster 1** | **Cluster 2** | **Cluster 3** | **Cluster 4** |
| **Cluster 1** | 76 | 0 | 3 | 0 |
| **Cluster 2** | 2 | 72 | 0 | 11 |
| **Cluster 3** | 1 | 1 | 53 | 2 |
| **Cluster 4** | 8 | 3 | 3 | 37 |

**Supplementary Figure 3: Results of Clustering When Fitting Mosquito Catch Data Using An Uninformative Prior.** In order to assess the sensitivity and robustness of the time series clustering, a less informative prior was used during the fitting process and the results displayed here. Top are the plots displaying the time series belonging to each cluster and which replicate the same 4 broad classes of temporal dynamics identified when clustering using the results from the fitting using an Informative Prior. Bottom table cross-tabulates Cluster assignments for individual time series across both sets of fitting – the majority (88%) of time series were consistently clustered across both sets of fitting, with the majority that displayed an incongruency being those belonging to Cluster 2 and Cluster 4 (from the Informative Prior fitting), the least peaked of the four temporal profiles. Predictive power based on the results of the multinomial logistic regression decreased when using the Uninformative Prior results, but remained substantially above that of a random classifier (predictive accuracy was 0.51, compared to the 0.25 expected for a truly random classifier and 0.58 for the model constructed using time series fitted with Informative Priors).

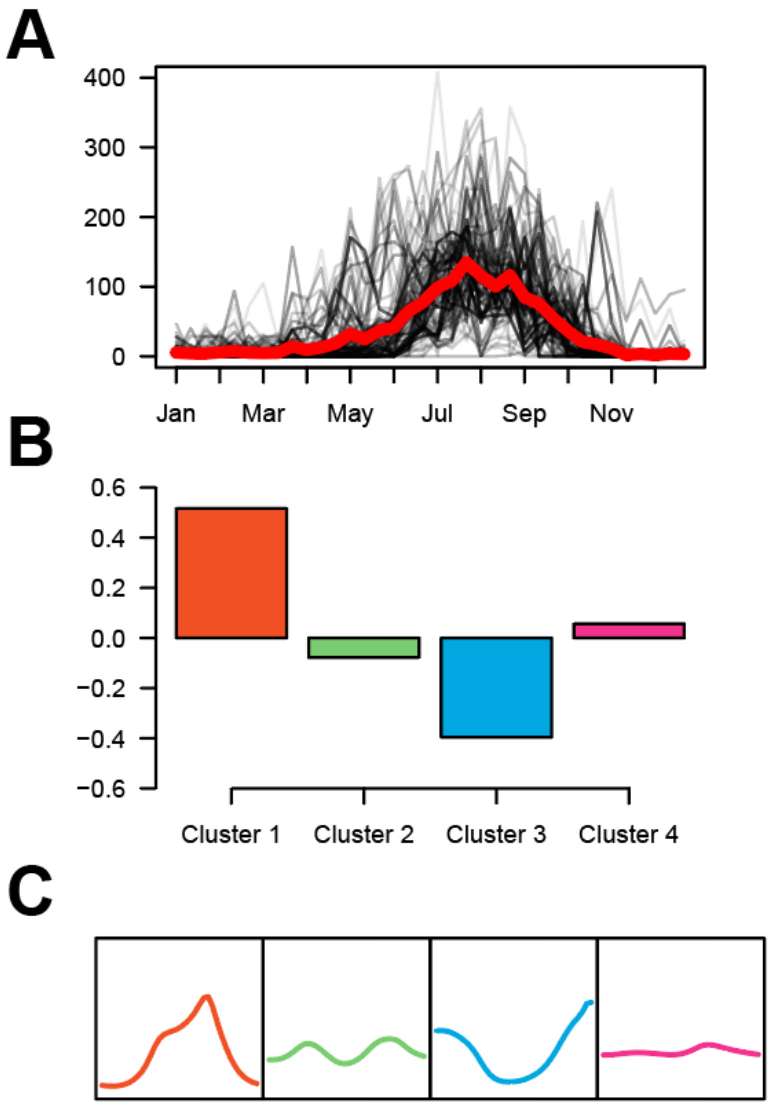


**Supplementary Figure 4: Exploring the Cross-Correlation Between Rainfall and Mosquito Densities.** For the 117 locations across India where mosquito catch data had been collected, daily, year specific rainfall data was also extracted and collated. This rainfall data was then aggregated up to the same temporal and spatial scale as the collected mosquito data and the cross-correlation between the two quantities explored. **(A)** Rainfall dynamics in India across the course of a year. Each black line represents the rainfall in a given location, whilst the thicker red line represents the average rainfall profile across all 117 locations. **(B)** Cluster specific cross-correlations between rainfall and mosquito catch size. **(C)** Mean mosquito catch temporal profiles for each Cluster.

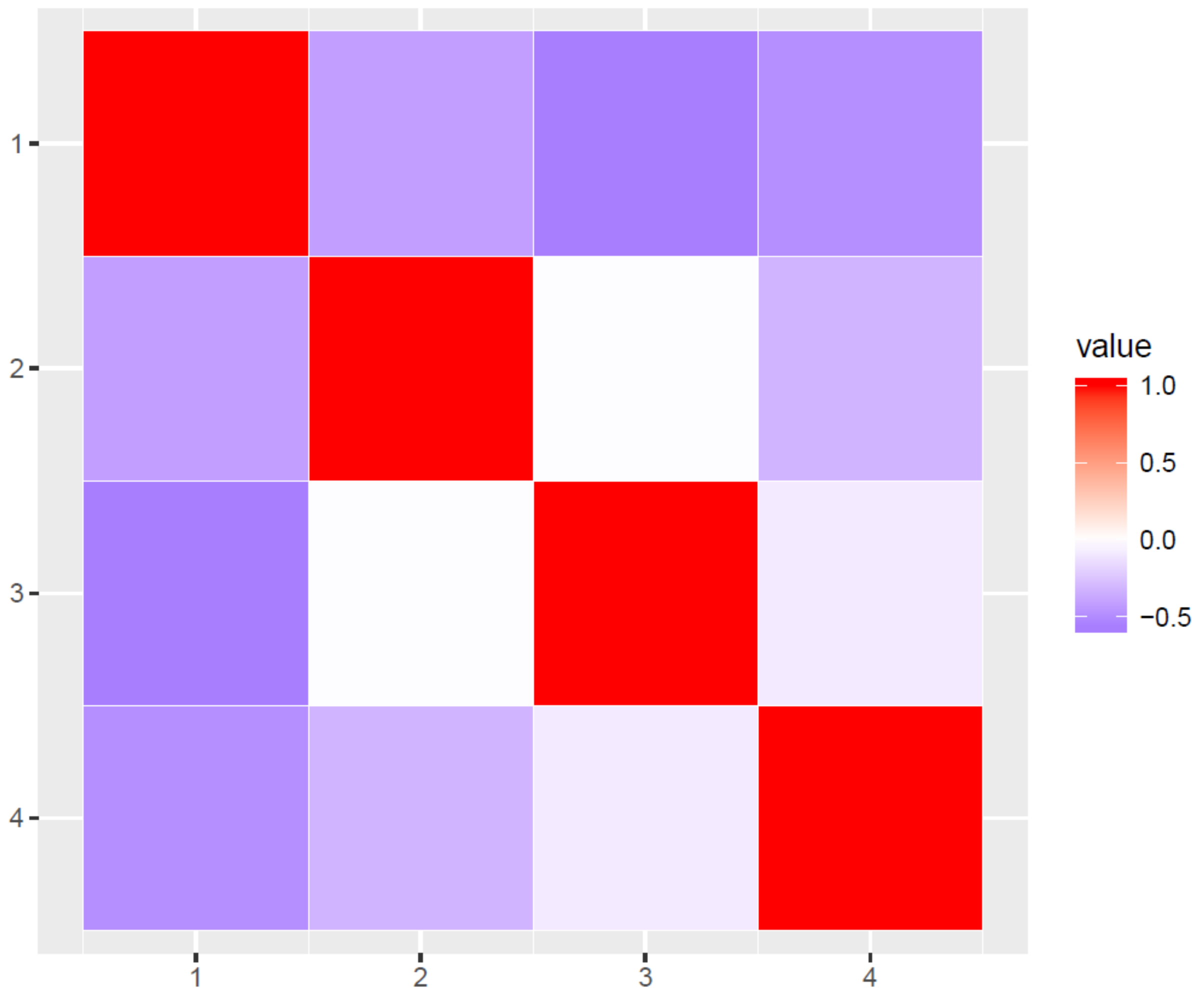


**Supplementary Figure 5: Cross-Cluster Correlations For Ecological Coefficients.** The cross-correlation between the predicted ecological coefficients from the multinomial logistic regression for each Cluster.
